# Social Determinants of Health and Cardiologist Involvement in the Care of Adults Hospitalized for Heart Failure

**DOI:** 10.1101/2023.03.23.23287671

**Authors:** David T. Zhang, Chukwuma Onyebeke, Musarrat Nahid, Lauren Balkan, Mahad Musse, Laura C. Pinheiro, Madeline R. Sterling, Raegan W. Durant, Todd M. Brown, Emily B. Levitan, Monika M. Safford, Parag Goyal

## Abstract

**Introduction:** The involvement of a cardiologist in the care of adults during a hospitalization for heart failure (HF) is associated with reduced rates of in-hospital mortality and hospital readmission. However, not all patients see a cardiologist when they are hospitalized for HF. Since reasons for this are not entirely clear, we sought to determine whether social determinants of health (SDOH) are associated with cardiologist involvement in the management of adults hospitalized for HF. We hypothesized that SDOH would be inversely associated with cardiologist involvement in the care of adults hospitalized for HF.

**Methods:** We included adult participants from the national REasons for Geographic And Racial Difference in Stroke (REGARDS) cohort, who experienced an adjudicated hospitalization for HF between 2009 and 2017. We excluded participants who were hospitalized at institutions that lacked cardiology services (n=246). We examined nine candidate SDOH, which align with the Healthy People 2030 conceptual model: Black race, social isolation (0–1 visits from a family or friend in the past month), social network/caregiver availability (having someone to care for them if ill), educational attainment < high school, annual household income < $35,000, living in rural areas, living in a zip code with high poverty, living in a Health Professional Shortage Area, and residing in a state with poor public health infrastructure. The primary outcome was cardiologist involvement, a binary variable which was defined as involvement of a cardiologist as the primary responsible clinician or as a consultant, collected via chart review. We examined associations between each SDOH and cardiologist involvement using Poisson regression with robust standard errors. Candidate SDOH with statistically significant associations (p<0.10) were retained for multivariable analysis. Potential confounders/covariates for the multivariable analysis included age, race, sex, HF characteristics, comorbidities, and hospital characteristics.

**Results:** We examined 876 participants hospitalized at 549 unique US hospitals. The median age was 77.5 years (IQR 71.0-83.7), 45.9% were female, 41.4% were Black, and 56.2% had low income. Low household income (<$35,000/year) was the only SDOH that had a statistically significant association with cardiologist involvement in a bivariate analysis (RR: 0.88 [95% CI: 0.82-0.95]). After adjusting for potential confounders, low income remained inversely associated (RR: 0.89 [95% CI: 0.82-0.97]).

**Conclusions:** Adults with low household income were 11% less likely to have a cardiologist involved in their care during a hospitalization for HF. This suggests that socioeconomic status may implicitly bias the care provided to patients hospitalized for HF.

## Introduction

Heart failure (HF) is the one of the most common reasons for hospitalization among adults, with high rates of readmission, healthcare costs, and mortality.^1-5^ The involvement of a cardiologist in the care of adults hospitalized for HF is associated with reduced rates of readmission and mortality.^6-16^ Yet, cardiologist involvement is not universal when adults are hospitalized for HF. Some of this might relate to cardiologist availability, which is especially problematic in rural settings plagued by shortages in cardiologists.^2, 7^

However, another potential contributor may be implicit bias. Indeed, in two studies of adults hospitalized with HF, Black race was associated with decreased odds of cardiologist involvement in the intensive care unit (ICU) and on the floor.^17, 18^ In another single-center study, both Black and Latinx adults hospitalized for HF were less likely to be admitted to a cardiology service and subsequently experienced worse outcomes.^19^ Differential utilization of cardiologists in the management of HF is not unique to race.^20-22^ In a multi-center study, Auerbach et al. (2000) found that adults admitted with severe HF who had an annual income less than $11,000 were less likely to receive care from a cardiologist.^17^ These observations raise concern that social determinants of health (SDOH), an increasingly recognized contributor to healthcare delivery,^23, 24^ lead to suboptimal care in the management of adults hospitalized for HF.

Notably, prior studies on this topic have not examined multiple SDOH concurrently and have lacked geographic diversity. With the current study, we sought to investigate the association between nine different candidate SDOH and the involvement of cardiologists in the care of adults hospitalized for HF, using the REasons for Geographic And Racial Differences in Stroke (REGARDS) cohort, which represents a geographically diverse cohort, includes HF hospitalizations from >500 unique hospitals, and offers access to a broad range of SDOH.^25-32^

## Methods

### Study Population

The study population included adults from REGARDS with an adjudicated HF hospitalization in 2009-2017. In brief, the REGARDS cohort originally included 30,239 community-dwelling Black and White men and women aged ≥45 years recruited via mailing followed by telephone contact in 2003-2007 from all 48 contiguous states in the United States and the District of Columbia to investigate racial and geographic disparities in stroke mortality.^33^ Black adults and residents of the Stroke Belt (Alabama, Arkansas, Georgia, Louisiana, Mississippi, North Carolina, South Carolina, and Tennessee), an area in the southeastern United States with high stroke mortality, were oversampled by design. Participants were recruited using commercially available lists. They underwent an initial 45-minute computer-assisted telephone interview about medical history, health behaviors, and risk factors followed by an in-home visit for baseline vitals, EKGs, medication reconciliation, bloodwork, and urine samples. At six-month intervals, participants were contacted by phone and asked about their health status and study outcomes such as hospitalizations. When participants report a hospitalization for a possible cardiovascular cause, their medical records were retrieved and underwent review and adjudication. HF adjudication was conducted by two expert clinicians based on chart-level data on symptoms, physical exams, laboratory values, imaging, and medical treatments. In the cases of a disagreement, a committee discussed to reach a consensus on a final decision.

For this study, we excluded participants hospitalized at institutions that lack cardiology services based on linked American Hospital Association data.^34^ Of note, there were some hospitals included in the American Hospital Association dataset that lacked information on the availability of cardiology services. To mitigate risk of bias related to excluded patients hospitalized at these hospitals, we imputed values for the availability of cardiology services using a random forest method that incorporated most other American Hospital Association data (including, but not limited to, location, bed size, teaching status, geriatrics service, palliative care service, licensed pharmacists). To do this, we used the R package “missForest.”^35, 36^ We examined each participant’s first adjudicated hospitalization during the study period.

The study protocol was reviewed and approved by Weill Cornell Medicine’s and the University of Alabama at Birmingham’s Institutional Review Boards. All participants provided written informed consent.

### Exposure: SDOH

The candidate exposures were nine SDOH available in REGARDS,^25-32^ following the Healthy People 2030 conceptual framework.^37^ This framework includes five domains: economic stability, education access and quality, social and community context, healthcare access and quality, and neighborhood and built environment.^37^ Figure 1 outlines the SDOH from REGARDS mapped onto the Healthy People 2030 conceptual framework.

**Figure 1:**
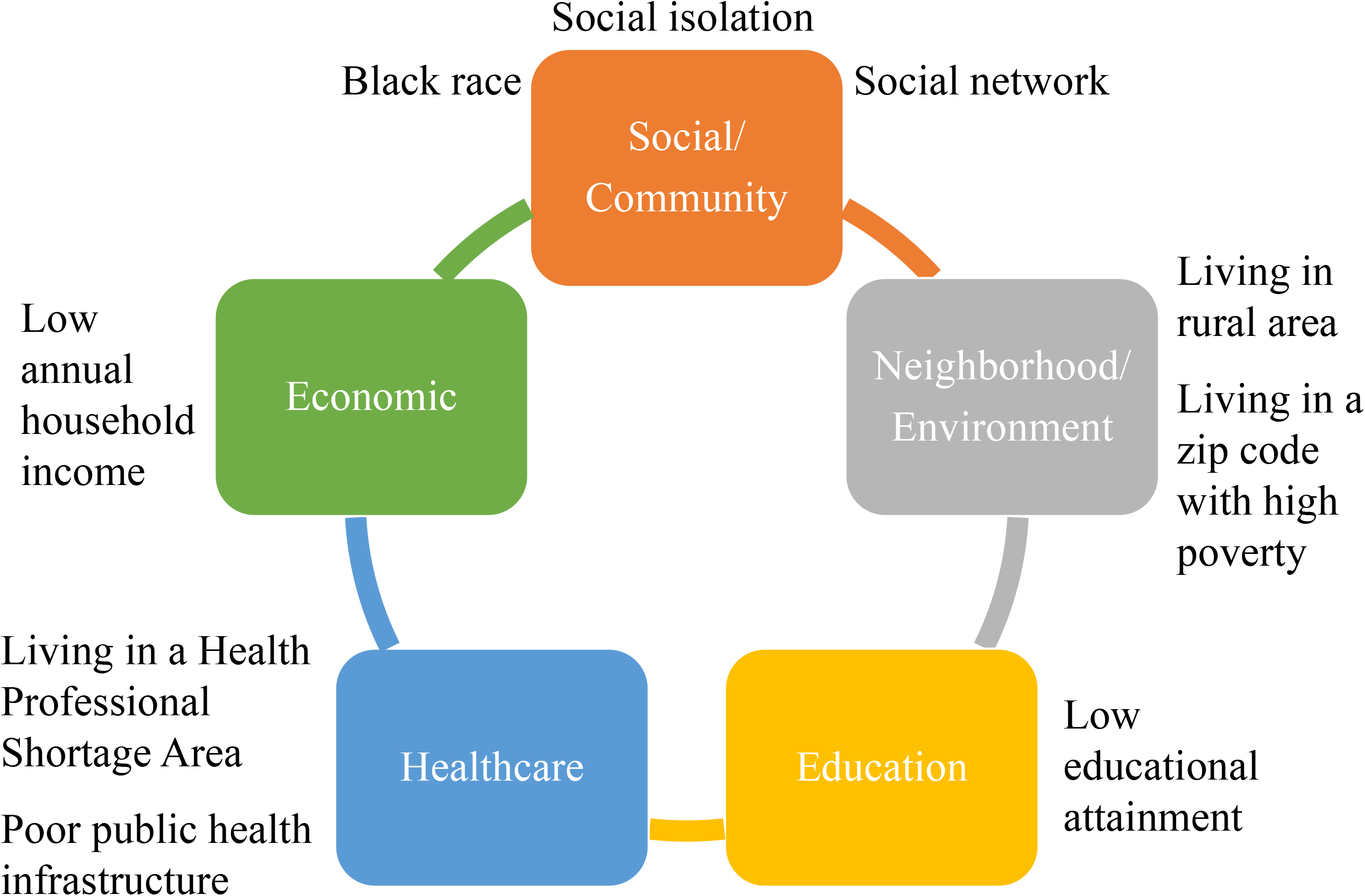
Healthy People 2030 Approach to Social Determinants of Health (SDOH)^37^ We studied nine SDOH, mapped onto the Healthy People 2030 conceptual framework (re-published from Sterling et al.,^25^ with permission).

Similar to prior work,^25-32^ we examined 1) Black race 2) social isolation (defined as having 0–1 visits from a family or friend in the past month), 3) social network/caregiver availability (defined as whether the participant reported having someone to care for them if ill), 4) low educational attainment (< high school education), 5) low annual household income (<$35,000/year), 6) living in rural areas (defined as living in an isolated or small rural area based off of Rural Urban Commuting Area Codes), 7) living in a zip code with high poverty (>25% of residence below the federal poverty line), 8) living in a Health Professional Shortage Area HPSA), and 9) living in a state with poor public health infrastructure (assessed using data from the America’s Health Ranking,^38^ which ranked states based on their contribution to lifestyle, access to care, and disability; states where REGARDS participants lived that fell into the bottom 20th percentile for their ranking for ≥8 of the 10 years spanning 2000-2010, concordant with REGARDS recruitment between 2003-2007, were considered to have poor public health infrastructure).

### Outcome

The primary outcome was cardiologist involvement, which was defined as involvement of a cardiologist as the primary responsible clinician or as a consultant. This variable was collected through a rigorous chart review and abstraction process previously described.^39^ Chart review and abstraction involved review of admission notes, progress notes, consultation notes, discharge summaries, medication reconciliations, laboratory values, and imaging studies to collect relevant data related to the HF hospitalization including cardiologist involvement.

### Covariates

Covariates were selected based on the Andersen Behavioral Model, which outlines predisposing, enabling, and need factors that contribute to healthcare utilization.^40^ Since the primary exposures (SDOH) are considered enabling factors, covariates included predisposing factors and need factors. Predisposing factors included age (at time of admission) and baseline self-reported race, sex, and an indicator for Stroke Belt residence. Need factors included the following three sub-categories: HF characteristics, comorbid conditions, and hospital characteristics—HF characteristics and comorbid conditions were collected based on chart abstraction, and hospital characteristics were based on American Hospital Association survey data which has been linked to the REGARDS cohort. HF characteristics included New York Heart Association Class (NYHA, Class I-IV), HF subtype (heart failure with preserved ejection fraction (HFpEF) if EF≥50% or qualitative description of normal systolic function),^41^ Get With The Guidelines-Heart Failure (GWTG-HF) Risk Score on admission (a score that predicts in-hospital all-cause mortality with a range of 0 to ≥79),^42^ length of stay, intensive care unit (ICU) stay, and in-hospital cardiac arrest. Comorbid conditions included coronary artery disease, history of arrythmia, diabetes mellitus, history of stroke, and the number of non-cardiac comorbid conditions (>40) spanning multiple organ systems. Hospital characteristics included each hospital’s total number of beds, presence/absence of cardiac ICU, and academic teaching status.

### Statistical Analysis

We calculated the frequency and distribution of each SDOH and examined collinearity using phi coefficients (Supplemental Table 1). We then examined associations between each SDOH and cardiologist involvement, using relative risks (RR). Similar to prior REGARDS studies, candidate SDOH with statistically significant associations (p<0.10) were retained for further multivariable analysis.^25-32^ All SDOH were collected at the time of initial REGARDS baseline surveys.

**Table 1:** Age-adjusted relative risks for the association of candidate SDOH with cardiologist involvement

| SDOH | Relative Risk (95% Confidence Intervals) | p-value |
| --- | --- | --- |
| Black race | 0.97 (0.90, 1.04) | 0.38 |
| Social isolation | 0.96 (0.86, 1.07) | 0.48 |
| Social network | 0.91 (0.80, 1.03) | 0.12 |
| Low educational attainment | 0.95 (0.86, 1.05) | 0.32 |
| Low annual household income | 0.88 (0.82, 0.95) | <0.001* |
| Living in rural areas | 0.98 (0.78, 1.23) | 0.85 |
| Living in a zip code with high poverty | 0.96 (0.88, 1.05) | 0.43 |
| Living in a Health Professional Shortage Area | 1.03 (0.96, 1.11) | 0.43 |
| Poor public health infrastructure | 1.03 (0.96, 1.11) | 0.39 |

\* retained for further analysis
SDOH = social determinants of health

To describe patient demographics, clinical, and hospital characteristics, descriptive statistics such as frequencies, percentages, medians, and inter-quartile ranges (IQR) were calculated. Chi-squared tests and Mann-Whitney tests were conducted for categorical and continuous variables, respectively, to identify significant differences (p<0.05) across categories of the retained SDOH.

We then examined the association between retained SDOH and cardiologist involvement by conducting a modified Poisson regression with robust standard error, adjusting for the previously described covariates. We chose a modified Poisson regression to estimate RR given the high prevalence of cardiologist involvement.^43^

We used two-sided hypothesis testing with p<0.05 for all analyses performed. Multiple imputation by chained equations was used to impute missing SDOH and covariates to minimize bias under the assumption that data were missing at random. All analyses were performed in Stata 17 (StataCorp, College Station, TX).

## Results

We examined 876 participants hospitalized at 549 unique US hospitals, of whom 75.3% received cardiology services while hospitalized. Figure S1 shows the exclusion cascade.

The prevalence of cardiologist involvement for each of the nine candidate SDOH are shown in Figure 2, and the associations between each of the SDOH and cardiologist involvement are shown in Table 1. Low income was the only candidate SDOH with a statistically significant association with cardiologist involvement: age-adjusted RR 0.88 (95% CI: 0.82-0.95, p<0.001) (Figure 3).

**Figure 2:**
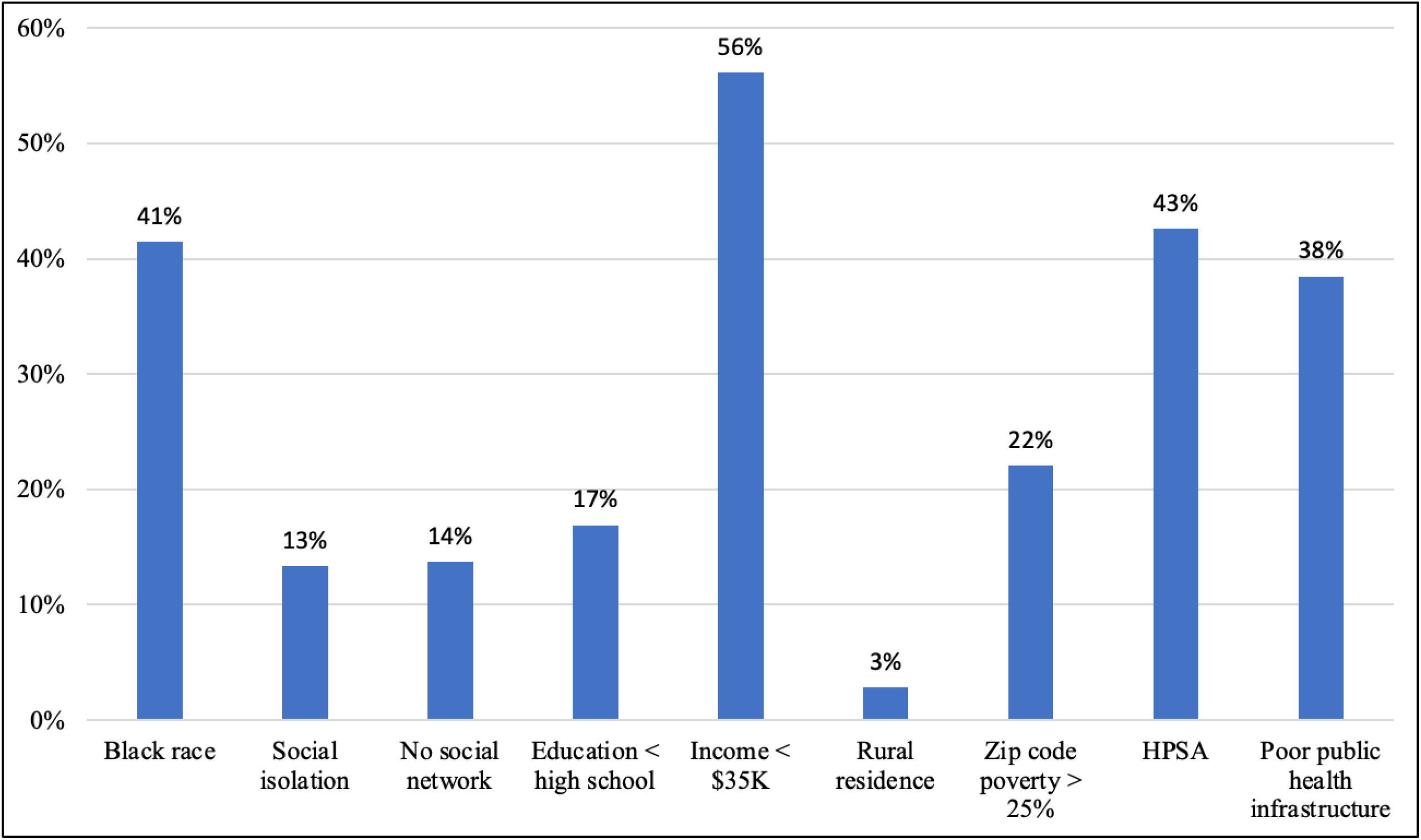
Prevalence of Individual SDOH The prevalence of the nine SDOH were calculated among REasons for Geographic And Racial Difference in Stroke (REGARDS) patients. Low annual household income (< $35,000/year) had the highest prevalence (56.2%) among SDOH.

**Figure 3:**
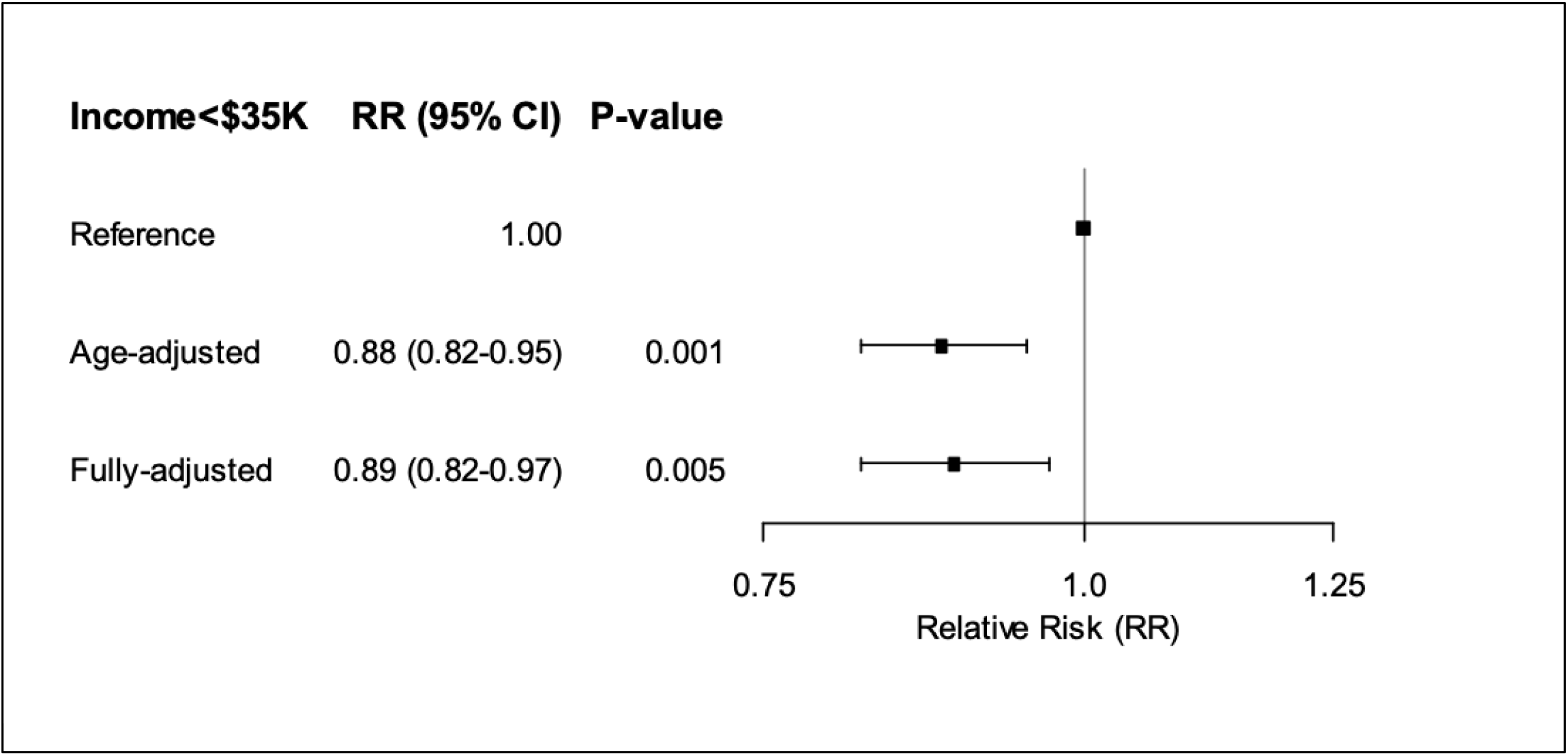
Relative Risk Ratios (RR) for Association Between Income <$35,000 and Cardiologist Involvement Low income was inversely associated with cardiologist involvement in both age-adjusted analysis (RR: 0.88 [95% CI: 0.82-0.95]) and fully-adjusted models (RR: 0.89 [95% CI: 0.82-0.97]).

Characteristics of the full cohort and participants stratified by income are shown in Table 2. The median age of the cohort was 77.5 years (IQR 71.0-83.7), 45.9% were female, 41.4% were Black, and 56.2% had low income. Participants with low income were more likely to be Black, female, from the Stroke Belt, and to have diabetes. Those with low income were more likely to have HFpEF, less likely to have a history of arrythmia, and had a lower GWTG-HF Risk Score on admission. Of note, 94.9% of the study cohort had insurance; and the prevalence was comparable among those who had and did not have involvement of a cardiologist in their care. The highest rates of missing data were HF subtype (24.2%), NYHA class (23.6%), presence of cardiac ICU (6.6%), and hospital teaching status (5.7%). All other covariates were missing <3%.

**Table 2:**
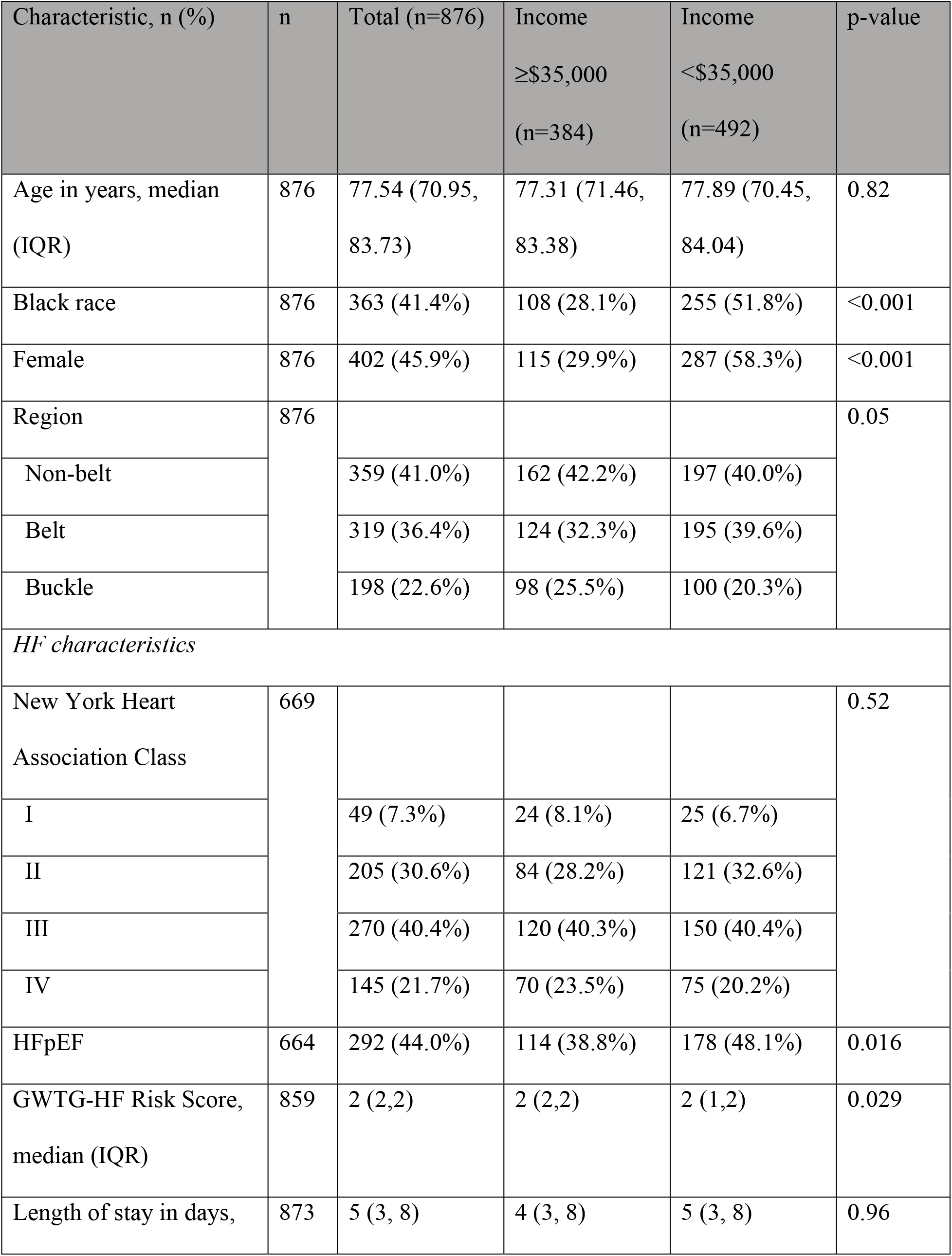
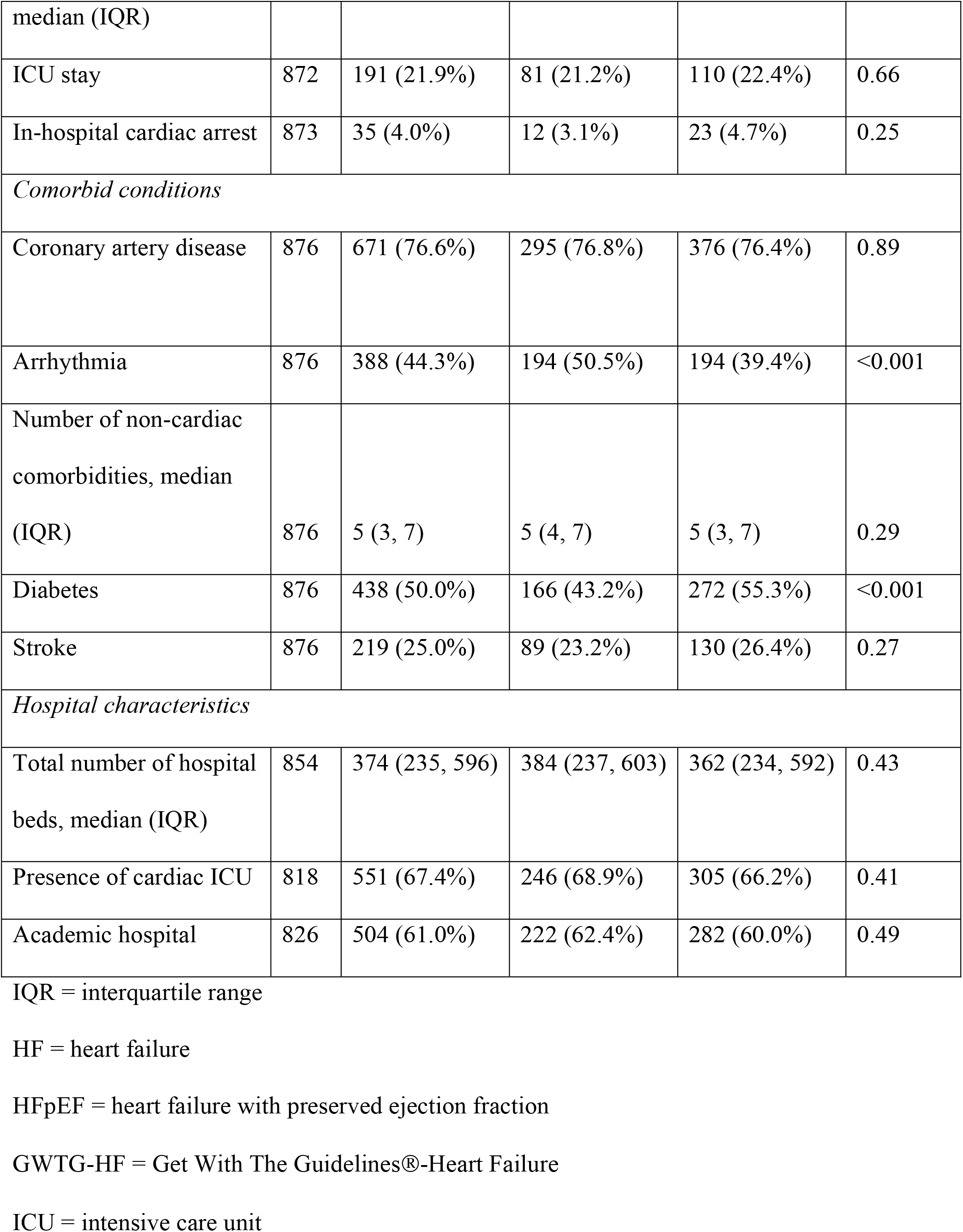
Participant characteristics stratified by annual household income

In a fully-adjusted model, low income remained inversely associated with cardiologist involvement during hospitalization (adjusted RR=0.89, 95% CI: 0.82-0.97, p<0.001) (Figure 3).

## Discussion

In this cohort study of adults hospitalized for HF, we found that low annual income (<$35,000/year) was associated with lower likelihood of cardiologist involvement during hospitalization. Prior work has shown the potential influence of SDOH on cardiologist involvement among adults with HF, but studies have been limited regarding the number and breadth of SDOH included. For example, Breathett et al. (2018) previously reported that Black race was inversely associated with receiving care from a cardiologist among adults admitted to the ICU with HF.^18^ This study included race, but did not include socioeconomic status (SES). Auerbach et al. (2000) showed that low income was inversely associated with having a cardiologist as the primary responsible clinician (primary attending);^17^ but their study only included those with advanced heart failure with reduced ejection fraction (HFrEF) and did not adjust for other potential SDOH like social isolation, social network, or area of residence. Our study included nine different SDOH (including race and income) that span multiple domains, and examined adults from >500 hospitals across the United States across the entire spectrum of left ventricular ejection fraction and severity. Accordingly, our study now provides the most comprehensive examination of SDOH and cardiologist involvement to date.

Our observations remained even after controlling for length of stay, severity of disease, HF subtype, and hospital characteristics. This raises concern that our observation could relate to implicit bias. Implicit bias is the subconscious stereotype or attitude that consequentially affects behavior and actions, which can contribute to healthcare disparities.^44, 45^ The concept of SDOH contributing to implicit bias with a subsequent effect on care provision is not new.^16, 46, 47^ However, this is one of the first studies to suggest that low income can contribute to implicit bias.^47^ Since income status may not be readily apparent to clinicians, further investigation is needed to better understand whether specific patient attributes or behaviors contribute to the suspected implicit bias observed among adults with low income. This phenomenon could relate to clinician perceptions about treatment adherence and/or affordability of services for adults with low income but will require further investigation. The vast majority of participants in this cohort had insurance; and we were unable to differentiate patients with different types of insurance.

Future work should examine whether insurance type is a contributor to the findings here. It is also not known whether lower rates of cardiologist involvement mediate the well-described associations between low socioeconomic status and hospital readmission or mortality among adults with HF.^48-51^ These findings should prompt further investigation into this possibility.

Our findings raise some questions related to a recent call for elicitation and documentation of SDOH experienced by adults with chronic diseases including HF.^44^ Given the ubiquitous role of the electronic medical record (EMR) in patient care, some have advocated for incorporating SDOH fields into the EMR, as a strategy to improve the care of vulnerable populations.^44, 52^ On the one hand, identifying adults with SDOH, such as low income, is important to identify a subpopulation at particularly high risk for poor outcomes. On the other hand, identification of SDOH could lead to implicit bias that negatively impacts care provision. Consequently, it is critical that efforts intended to increase elicitation and documentation of SDOH be paired with strategies to address implicit bias from the healthcare system. Some previously described strategies to address implicit bias among clinicians in the healthcare system include implicit bias training, increasing diversity among healthcare workers, and partnering with community advocates.^44, 45, 53-57^ Clearly, effort is needed to routinely incorporate these strategies as a means to address the multi-tiered effects of SDOH.

Our study has several strengths. First, we examined a geographically diverse cohort that included data on multiple (nine) SDOH that map onto an existing Healthy People 2030 framework.^37^ In addition, we had chart-level data at the time of the HF hospitalization, which included multiple factors that could have impacted cardiologist involvement—this allowed us to adjust for several important confounders including HF severity. There are also some important limitations to note. First, the study sample included White and Black participants, but did not include other minoritized populations. Future studies are needed to understand the impact of SDOH on these groups. Second, although we examined nine different SDOH, we did not conduct a detailed examination of other relevant factors like health literacy, medication adherence, health insurance, or frailty. Finally, although we had chart-level data, involvement of a cardiologist may not have been clear in all scenarios, which could have led to misclassification bias.

## Conclusion

We found that low income was inversely associated with cardiologist involvement during hospitalization after adjusting for potential confounders. This suggests that certain social factors may implicitly bias the care provided to patients hospitalized for HF, calling attention to a potentially important source of disparity in care.

## Data Availability

The authors thank the other investigators, the staff, and the participants of the REGARDS study (REasons for Geographic And Racial Differences in Stroke) for their valuable contributions. A full list of participating REGARDS investigators and institutions can be found at http://www.regardsstudy.org.

---

**Supplemental Figure 1:**
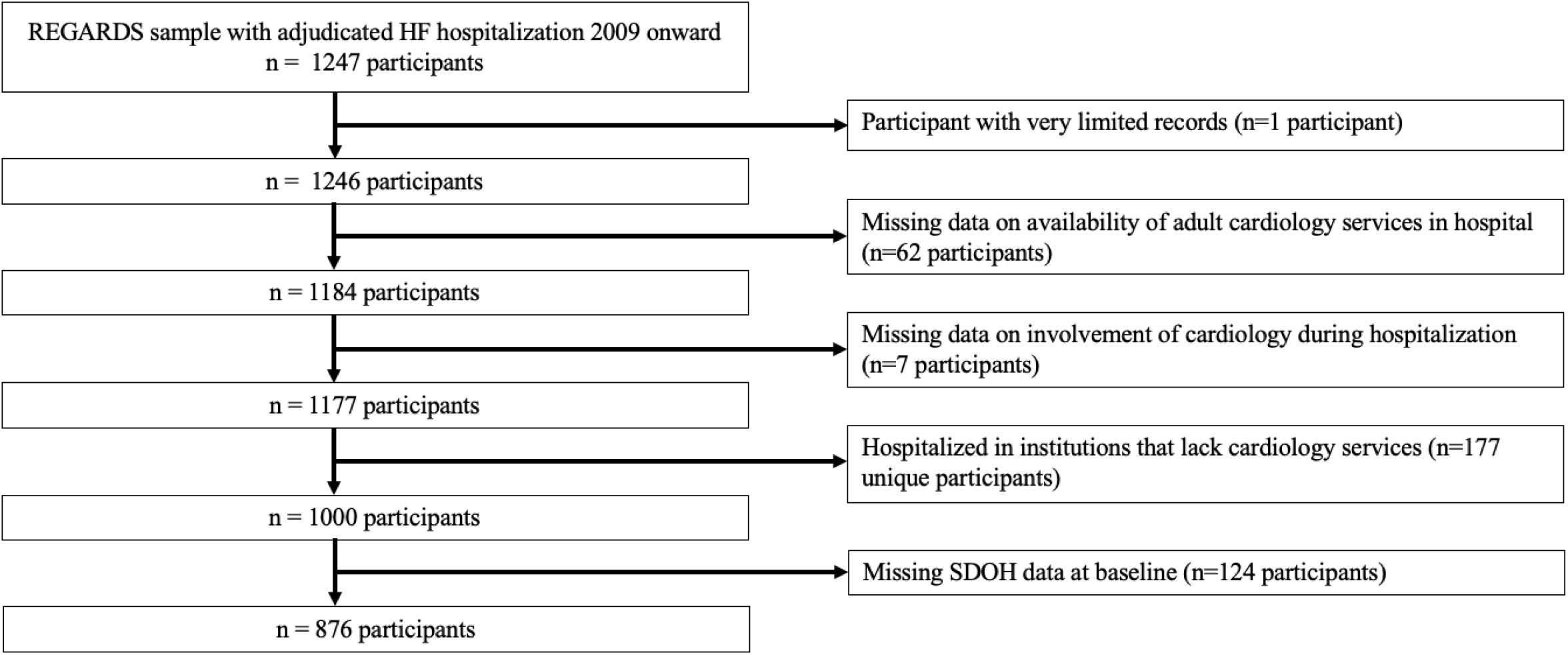
Inclusion/Exclusion Cascade

